# Transmitted HIV-1 is more pathogenic in heterosexual individuals than homosexual men

**DOI:** 10.1101/2020.09.08.20191015

**Authors:** Ananthu James, Narendra M. Dixit

**Affiliations:** Department of Chemical Engineering, Indian Institute of Science, Bengaluru, India; Centre for Biosystems Science and Engineering, Indian Institute of Science, Bengaluru, India

## Abstract

Transmission bottlenecks introduce selection pressures on HIV-1 that vary substantially with the mode of transmission. Recent studies on small cohorts have suggested that stronger selection pressures lead to fitter transmitted/founder (T/F) strains. Manifestations of this selection bias at the population level have remained elusive. Here, we analysed early CD4 cell count measurements reported from ∼340,000 infected heterosexual individuals (HSX) and men-who-have-sex-with-men (MSM), across geographies, ethnicities and calendar years and found them to be consistently lower in HSX than MSM (P<0.05). The corresponding average reduction in CD4 counts relative to healthy adults was 86.5% in HSX and 67.8% in MSM (P<10^−4^). This difference could not be attributed to differences in age, HIV-1 subtype, viral load, gender, ethnicity, time of transmission, or diagnosis delay across the groups. We concluded that the different selection pressures arising from the different predominant transmission modes have resulted in more pathogenic T/F strains in HSX than MSM.

## Introduction

The bottlenecks in HIV-1 transmission result in a ‘selection bias’ favoring fitter transmitted/founder (T/F) viruses over less fit ones^1,2^. Several recent studies have presented evidence of genetic, phenotypic, and clinical manifestations of the selection bias in small cohorts^1,3–6^. From 137 heterosexual (HSX) donor-recipient pairs, T/F viruses were found to carry higher than average frequencies of amino acids associated with high *in vivo* fitness^1^. Similarly, from 127 discordant couples, lower viral replication capacity (vRC), indicative of lower viral fitness, early in infection was associated with slower decline of CD4 T cell counts^4,6^. The selection bias varies with the mode of transmission^3^. The stronger the bottlenecks, the fitter the corresponding T/F viruses are likely to be^1,2^. Anal intercourse is over 10-fold more permissive on average than penile-vaginal intercourse^7^. Analysis of T/F genomes from 131 subjects revealed that the T/F genomes were under greater positive selection in heterosexual individuals (HSX), in whom the penile-vaginal mode predominates^8^, than homosexual men, or men-who-have-sex-with-men (MSM), who transmit predominantly through anal intercourse^3^. Among HSX, men had T/F viruses with higher predicted fitness *in vivo* than women^1^, consistent with the asymmetry of the bottlenecks between insertive and receptive penile-vaginal intercourse^7^.

An important question that follows is whether the differential selection bias across modes of transmission is manifested at the wider population level. Such differential bias could contribute to variations in disease progression and treatment outcomes and underlie the diverse trajectories of the HIV-1 pandemic across infected groups in which different modes of transmission predominate.

## Results and Discussion

To answer this question, we decided to compare early CD4 T cell count measurements between HSX and MSM. Immediately following infection, CD4 T cell counts fall steeply, recover partially, and then settle within a few weeks/months to a value smaller than in the pre-infection state^9^(Fig. 1(a)). Subsequent changes in the CD4 counts occur slowly, over many months to years. Thus, CD4 count measurements made early in infection tend to be close to the value to which the counts settle after the initial dynamics. These early CD4 counts are expected to be minimally affected by host-specific adaptive mutations^1^ and, therefore, representative of the fitness of the T/F strain in the recipient. The fitter the strain, the lower would be the CD4 count. The CD4 count is also a more robust marker of disease state than other commonly used markers such as set-point viral load (SPVL). High vRC of the T/F viruses was associated with low CD4 counts at 3 months post-infection (which roughly coincides with seroconversion) and rapid CD4 count decline for ∼5 years, independently of SPVL^4,6^.

**Figure 1:**
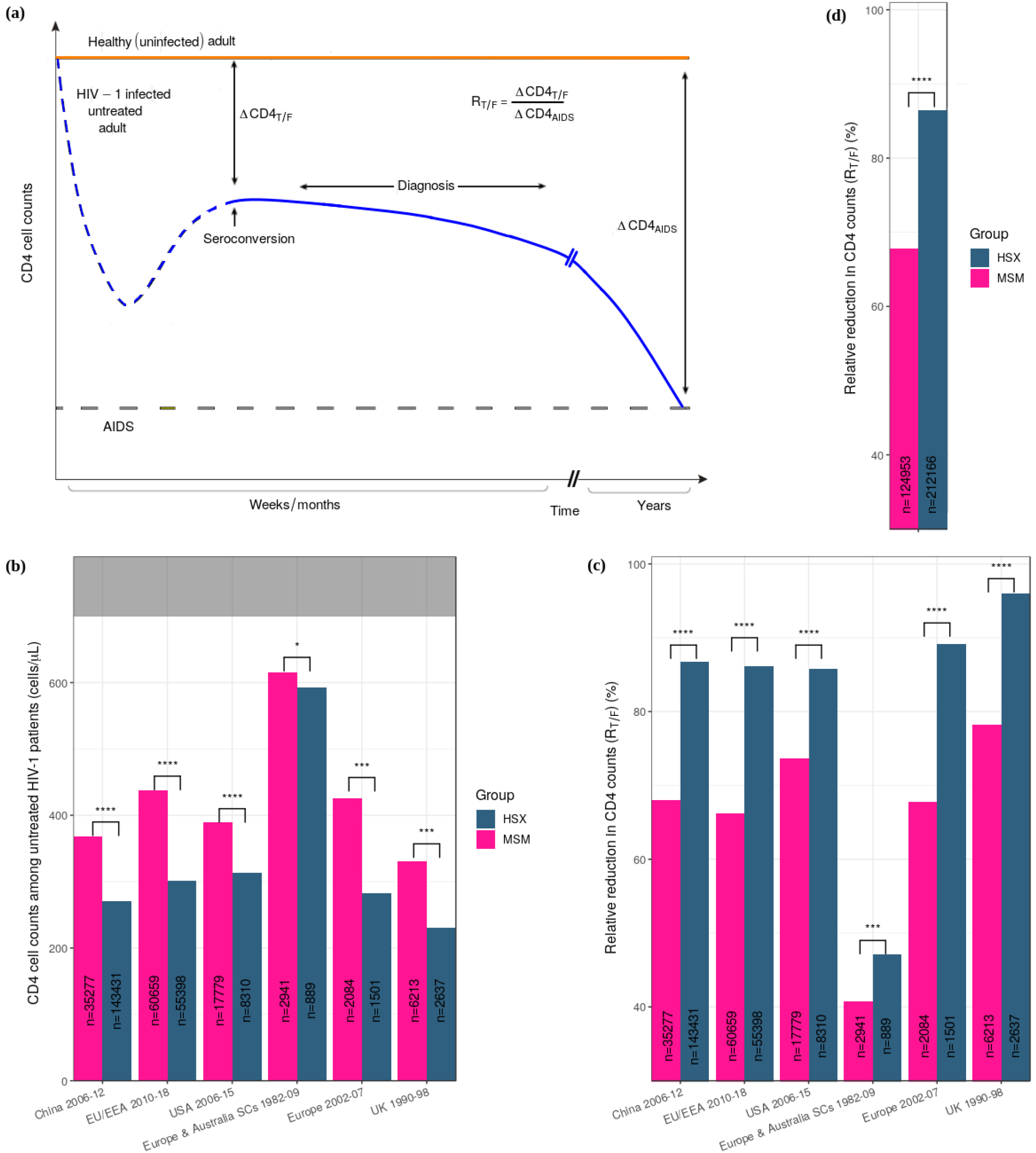
Early CD4 T cell counts and the associated relative reduction (*R_T/F_*) in MSM and HSX. **(a)** Schematic of typical CD4 count changes post HIV-1 infection (blue), before (dashed) and after (solid) diagnosis/seroconversion. The reduction at diagnosis/seroconversion relative to uninfected individuals (orange) and that associated with AIDS (grey dashed line) yields *R_T/F_*, the reduction attributable to the T/F virus. **(b)** Early mean CD4 cell counts and **(c)** the corresponding *R_T/F_* in untreated infected adult HSX and MSM from different geographical regions and calendar years (see Methods, Tables 1 and S3-S5 for details). The grey region indicates counts in uninfected, healthy individuals. **(d)** Population-weighted average of *R_T/F_* across all the datasets in (c). The sample sizes (*n*) are indicated. SCs indicate seroconverters. ****, ***, ** and * indicate P< 10^−4^, P< 10^−3^, P< 10^−2^and P< 0.05, respectively.

HSX and MSM are the two major groups driving the global HIV-1 epidemic^9^. They use predominant modes of transmission with a substantial difference in the selection bias^7^. Importantly, they display little inter-mixing in most geographical regions. We inferred the latter from the distinct prevalence of HIV-1 subtypes in the two groups, which we found across geographical regions and calendar years (Fig. 2; Text S1; Tables S1 and S2). Together, these characteristics allow for the difference in the selection bias to be sustained long-term, potentially amplified, and manifested in sample sizes large enough for detection with statistical significance. We thus hypothesized that the stronger selection bias associated with penile-vaginal transmission than anal transmission would result in lower early CD4 counts in HSX than in MSM.

**Figure 2:**
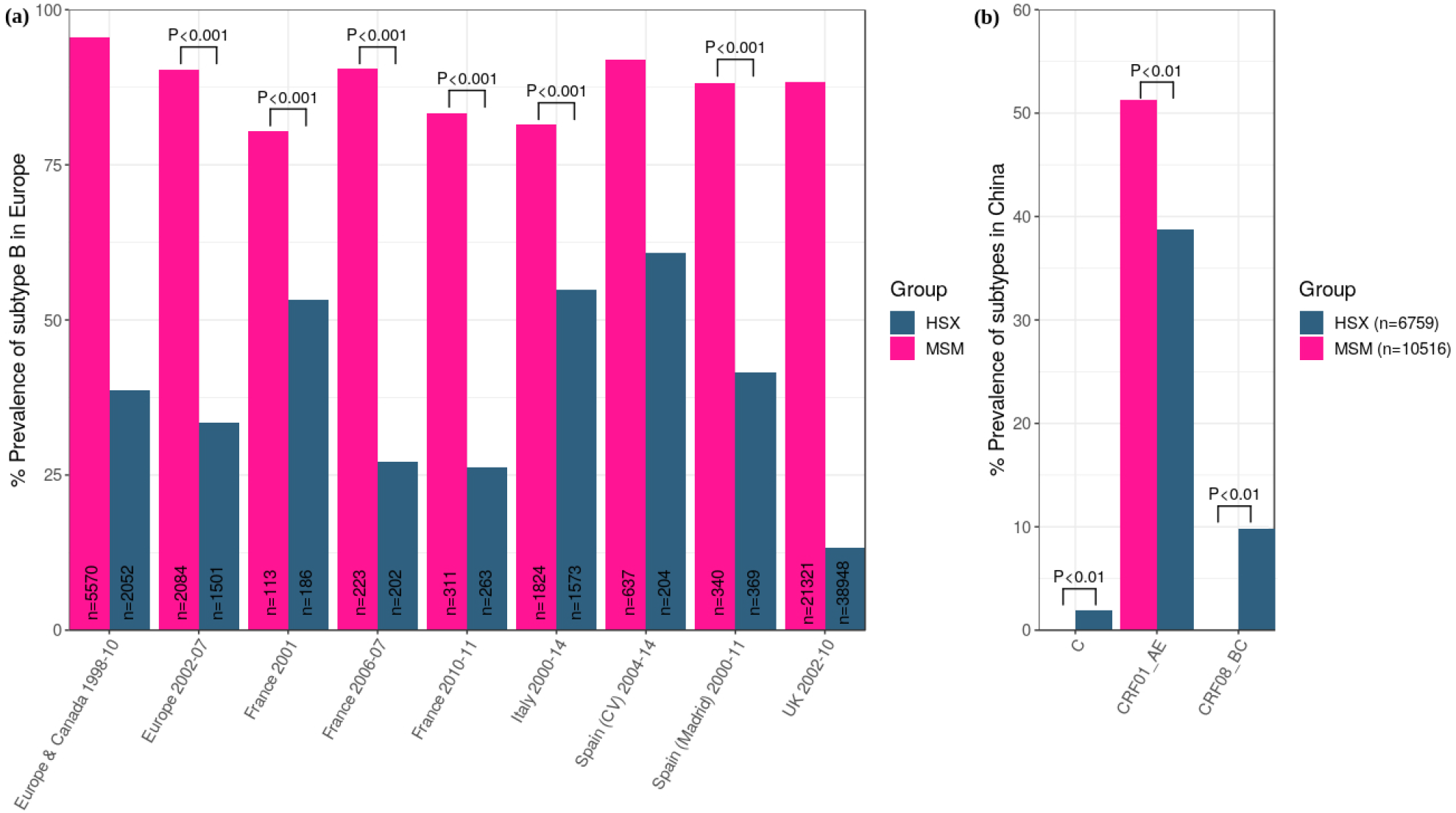
Subtype prevalence of HIV-1 in MSM and HSX populations. Prevalence of **(a)** subtype B in different regions in Europe and Canada and **(b)** all the subtypes in China. The sample sizes (*n*) along with the time periods of the surveys are indicated. *P* values are listed where available in the original sources. Sources of the data and additional details are in Tables S1 and S2. The different prevalence indicates little mixing between MSM and HSX in the populations studied.

To test this hypothesis, we collated available data of CD4 count measurements either at seroconversion or at diagnosis from all large studies, which amounted to a total of ∼340,000 patients across four geographical regions followed over a total period of nearly four decades, and examined the differences between HSX and MSM (Methods; Table 1). We found that HSX consistently had lower CD4 counts than MSM (Fig. 1(b); Tables 1 and Tables S3-S5). For instance, measurements from ∼120,000 patients across 21 countries in the European Union and European Economic Area (EU/EEA) indicated, following population-weighted averaging of yearly data during 2010–2018, that the mean CD4 count in MSM at diagnosis was ∼440 cells/*μ*L, whereas it was substantially lower, ∼300 cells/*μ*L, in HSX (P<10^−4^)^10^. The numbers were similar in the preceding 5 year period (2002–2007) reported by a smaller study involving a few thousand patients^11^. In the UK, measurements from close to 9000 patients during 1990–1998 showed that the counts at diagnosis were ∼330 cells/*μ*L in MSM and ∼230 cells/*μ*L in HSX (P<10^−3^)^12^. In China, during 2006–2012, the mean CD4 counts at diagnosis from ∼180,000 patients were ∼370 cells/*μ*L in MSM and ∼270 cells/*μ*L in HSX (P<10^−4^)^13^. Similarly, in the US, from over 25,000 patients during 2006–2015, the counts at diagnosis were ∼400 cells/*μ*L in MSM and ∼300 cells/*μ*L in HSX (P<10^−4^)^14^. We also examined/estimated the counts at seroconversion where available. In the CASCADE study, involving ∼4000 patients during 1979–2000 in Europe and Australia, the mean cell counts at seroconversion were ∼620 cells/*μ*L in MSM and ∼590 cells/*μ*L in HSX (P = 0.027)^15^. Further, using the reported diagnosis delays and the slopes of CD4 count decline in the US population above^14^, we estimated that the cell counts at seroconversion, for the age group 13–29 years, were ∼550 cells/*μ*L in MSM and ∼480 cells/*μ*L in HSX (P<10^−4^) (Methods). Remarkably, we did not find any large study (sample size & 1000) that reported higher early CD4 cell counts in HSX than MSM.

**Table 1.**
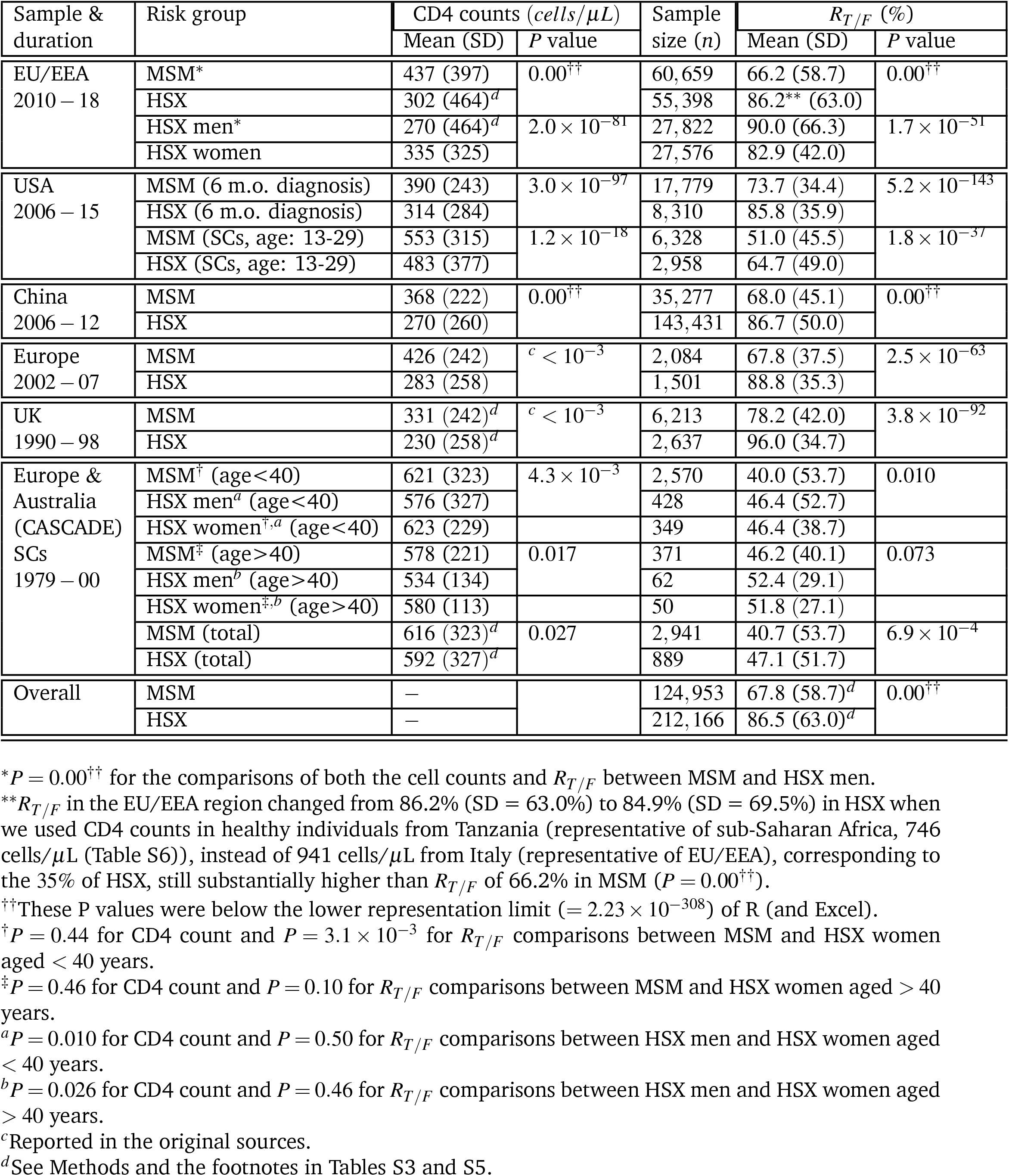
Early CD4 cell counts in infected adults at diagnosis or seroconversion, estimated from data collated from several studies (see text and Tables S3-S5). The corresponding relative reduction in CD4 count, *R_T/F_*, calculated as described in the text, are also listed. *P* values for comparisons of the CD4 counts (and *R_T/F_*) between MSM and HSX indicate a significantly higher CD4 counts and lower *R_T/F_* in MSM throughout. The last row represents the population-weighted average of all the datasets.

| Sample & duration | Risk group | CD4 counts (*cells/μL*) | | Sample size (*n*) | $R_{T/F}$ (%) | |
| --- | --- | --- | --- | --- | --- | --- |
| | | Mean (SD) | $P$ value | | Mean (SD) | $P$ value |
| EU/EEA<br>2010 – 18 | MSM* | 437 (397) | 0.00<sup>††</sup> | 60,659 | 66.2 (58.7) | 0.00<sup>††</sup> |
|  | HSX | 302 (464)<sup>d</sup> |  | 55,398 | 86.2** (63.0) |  |
| | HSX men* | 270 (464)<sup>d</sup> | $2.0 \times 10^{-81}$ | 27,822 | 90.0 (66.3) | $1.7 \times 10^{-51}$ |
|  | HSX women | 335 (325) |  | 27,576 | 82.9 (42.0) |  |
| USA<br>2006 – 15 | MSM (6 m.o. diagnosis) | 390 (243) | $3.0 \times 10^{-97}$ | 17,779 | 73.7 (34.4) | $5.2 \times 10^{-143}$ |
|  | HSX (6 m.o. diagnosis) | 314 (284) |  | 8,310 | 85.8 (35.9) |  |
| | MSM (SCs, age: 13-29) | 553 (315) | $1.2 \times 10^{-18}$ | 6,328 | 51.0 (45.5) | $1.8 \times 10^{-37}$ |
|  | HSX (SCs, age: 13-29) | 483 (377) |  | 2,958 | 64.7 (49.0) |  |
| China<br>2006 – 12 | MSM | 368 (222) | 0.00<sup>††</sup> | 35,277 | 68.0 (45.1) | 0.00<sup>††</sup> |
|  | HSX | 270 (260) |  | 143,431 | 86.7 (50.0) |  |
| Europe<br>2002 – 07 | MSM | 426 (242) | $c < 10^{-3}$ | 2,084 | 67.8 (37.5) | $2.5 \times 10^{-63}$ |
|  | HSX | 283 (258) |  | 1,501 | 88.8 (35.3) |  |
| UK<br>1990 – 98 | MSM | 331 (242)<sup>d</sup> | $c < 10^{-3}$ | 6,213 | 78.2 (42.0) | $3.8 \times 10^{-92}$ |
|  | HSX | 230 (258)<sup>d</sup> |  | 2,637 | 96.0 (34.7) |  |
| Europe & Australia<br>(CASCADE)<br>SCs<br>1979 – 00 | MSM<sup>†</sup> (age<40) | 621 (323) | $4.3 \times 10^{-3}$ | 2,570 | 40.0 (53.7) | 0.010 |
|  | HSX men<sup>a</sup> (age<40) | 576 (327) |  | 428 | 46.4 (52.7) |  |
|  | HSX women<sup>†,a</sup> (age<40) | 623 (229) |  | 349 | 46.4 (38.7) |  |
|  | MSM<sup>‡</sup> (age>40) | 578 (221) | 0.017 | 371 | 46.2 (40.1) | 0.073 |
|  | HSX men<sup>b</sup> (age>40) | 534 (134) |  | 62 | 52.4 (29.1) |  |
|  | HSX women<sup>‡,b</sup> (age>40) | 580 (113) |  | 50 | 51.8 (27.1) |  |
| | MSM (total) | 616 (323)<sup>d</sup> | 0.027 | 2,941 | 40.7 (53.7) | $6.9 \times 10^{-4}$ |
|  | HSX (total) | 592 (327)<sup>d</sup> |  | 889 | 47.1 (51.7) |  |
| Overall | MSM | — |  | 124,953 | 67.8 (58.7)<sup>d</sup> | 0.00<sup>††</sup> |
|  | HSX | — |  | 212,166 | 86.5 (63.0)<sup>d</sup> |  |

\* $P = 0.00^{\dagger\dagger}$ for the comparisons of both the cell counts and $R_{T/F}$ between MSM and HSX men.
\*\* $R_{T/F}$ in the EU/EEA region changed from 86.2% (SD = 63.0%) to 84.9% (SD = 69.5%) in HSX when we used CD4 counts in healthy individuals from Tanzania (representative of sub-Saharan Africa, 746 cells/μL (Table S6)), instead of 941 cells/μL from Italy (representative of EU/EEA), corresponding to the 35% of HSX, still substantially higher than $R_{T/F}$ of 66.2% in MSM ( $P = 0.00^{\dagger\dagger}$ ).
<sup>††</sup>These $P$ values were below the lower representation limit ( $= 2.23 \times 10^{-308}$ ) of R (and Excel).
<sup>†</sup> $P = 0.44$ for CD4 count and $P = 3.1 \times 10^{-3}$ for $R_{T/F}$ comparisons between MSM and HSX women aged < 40 years.
<sup>‡</sup> $P = 0.46$ for CD4 count and $P = 0.10$ for $R_{T/F}$ comparisons between MSM and HSX women aged > 40 years.
<sup>a</sup> $P = 0.010$ for CD4 count and $P = 0.50$ for $R_{T/F}$ comparisons between HSX men and HSX women aged < 40 years.
<sup>b</sup> $P = 0.026$ for CD4 count and $P = 0.46$ for $R_{T/F}$ comparisons between HSX men and HSX women aged > 40 years.
<sup>c</sup>Reported in the original sources.
<sup>d</sup>See Methods and the footnotes in Tables S3 and S5.

While the evidence from absolute CD4 count comparisons was thus overwhelming, differences in CD4 counts in healthy (uninfected) individuals across gender, ethnicity and geographical regions could render absolute CD4 counts only an approximate measure of the fitness of the T/F strains. Two individuals may have similar early CD4 counts but may still have been infected by T/F strains of different fitness if their pre-infection CD4 counts were different, with the individual with the higher pre-infection count infected by the fitter T/F strain. To overcome this limitation, we constructed a metric to quantify the relative reduction in the CD4 cell count, *R*, corresponding to the absolute CD4 count *T* as 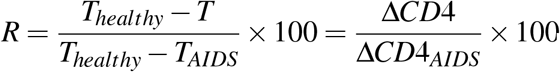, where *T_healthy_* was the count pre-infection, and *T_AIDS_* = 200 cells/*μ*L the count defining AIDS. Thus, *R* was 0% when *T* = *T_healthy_* and 100% when *T* = *T_AIDS_* and decreased linearly with *T* between these extremes. Choosing *T_healthy_* specific to the respective geographies, ethnicities, and genders (Table S6), we estimated *R* corresponding to the early cell count measurements above, which we denoted as *R_T/F_*, indicative of the relative reduction in CD4 count due to the T/F virus (Fig. 1(c)). The higher the *R_T/F_*, the fitter would be the T/F strain, regardless of the pre-infection CD4 count, rendering *R_T/F_* a more robust marker of T/F viral fitness than the associated early absolute CD4 counts. (Note that *R_T/F_* is a static measure and is not indicative of the ‘speed’ of disease progression; cell count decline can be faster despite higher early CD4 counts in MSM than HSX^15,16^.)

We found that in EU/EAA, during 2010–18, *R_T/F_* was 86.2% in HSX and 66.2% in MSM (P<10^−4^). During 2002–07, these numbers were 88.8% and 67.8% (P<10^−4^), respectively. The corresponding numbers were 96.0% and 78.2% in the UK (P<10^−4^), and 86.7% and 68.0% in China (P<10^−4^).

In the US, the difference was smaller but still substantial, with *R_T/F_* of 85.8% in HSX and 73.7% in MSM (P<10^−4^). At seroconversion, these numbers were 64.7% and 51.0%, respectively (P<10^−4^). For the seroconverters from the CASCADE study, the trend was consistent, with *R_T/F_* of 47.1% in HSX and 40.7% in MSM (P<10^−3^). Overall, thus, *R_T/F_* comparisons showed more significant differences between MSM and HSX than absolute CD4 count comparisons (Fig. 1(b) and (c)). Further, *R_T/_F* allowed comparison across the different datasets. Thus, while the HSX all had *R_T/F_* >85% at diagnosis, the MSM displayed a range from ∼65% to a little under 80%. We could also combine the datasets, including those at diagnosis and seroconversion, and estimate an overall *R_T/F_*. Using a population-weighted average across the datasets, we estimated the overall *R_T/F_* to be 86.5% in HSX and 67.8% in MSM (P<10^−4^) (Fig. 1(d)). This overall comparison provides strong evidence of greater cell count reduction due to, and hence greater pathogenicity of, the T/F viruses in HSX than in MSM.

To attribute the differences in *R_T/F_* between HSX and MSM to the differential selection bias at transmission in the two groups, we considered and ruled out all the major potential confounding factors. First, MSM are typically diagnosed at a younger age than HSX. In the two European studies, MSM were 5 (Table S5)^10^ and 1.6 years^11^ younger on average than HSX at diagnosis. Given the cell count decrease of ∼7 cells/*μ*L per year of age at diagnosis^12^, the CD4 counts should have been higher in MSM by only ∼35 and ∼11 cells/*μ*L, whereas they were higher by 135 and 143 cells/*μ*L (Fig. 1(b)), respectively, a difference that could not be explained by the age at diagnosis. Second, MSM are often predominantly infected by subtype B^17^, whereas HSX are by subtypes B and C (Fig. 2; Text S1). This subtype difference should have resulted in lower CD4 counts in MSM than HSX because of the higher virulence of subtype B^18,19^, a trend opposite of what is observed. Moreover, in the US where subtype B dominates both HSX and MSM (Text S1), *R_T/F_* was lower among MSM (Fig. 1(c)). In agreement, an independent study found that subtype B T/F viruses had higher fitness among HSX than MSM^3^.

Third, the CD4 counts could not be explained as an indirect manifestation of variations in SPVL; in the European study, CD4 counts were higher in MSM despite higher SPVL in MSM than HSX (Table S3). Fourth, healthy men had lower CD4 counts than HSX and healthy women everywhere except China (Table S6), and infected HSX men displayed higher *R_T/F_* than MSM (Table 1 and Fig. S1), two reasons to rule out gender as the cause of lower *R_T/F_* in MSM. Fifth, in Europe (EU/EEA), while MSM are predominantly Caucasian, 30–35% of infected HSX are of sub-Saharan African origin^10,11^.

In China, however, where no differences in ethnicity exist between MSM and HSX, a substantial difference in *R_T/F_* is seen between them (Fig. 1(c)), ruling out ethnicity as a confounding factor.

Further, accounting for baseline CD4 count differences across ethnicities in EU/EEA did not alter our findings (Table 1). Sixth, early onward transmission may limit donor-specific adaptations in the T/F strain and allow it to cause more severe cell count reduction in the recipient. Early transmissions, however, are more common to MSM than HSX^18,20^, in keeping with the greater association of MSM with transmission clusters^17^(Fig. 3; Table S7), and should have led to higher *R_T/F_* in MSM than HSX, in contrast to our findings. Seventh, although MSM tend to be diagnosed earlier than HSX^14^ and may thus suffer a lower loss of CD4 counts at diagnosis, the differences are seen also in CD4 counts at seroconversion^14,15^, which would occur at similar times post infection in the two groups. Besides, MSM had lower cell counts in China too, where, owing to social stigma, MSM may not get diagnosed earlier than HSX^13^. The difference in *R_T/F_* between MSM and HSX was thus not attributable to any of the above factors. We concluded therefore that the difference originated from the variations in the fitness of the T/F strains in the two groups arising from the different selection biases at transmission.

Our findings establish the selection bias at transmission as an important underlying factor shaping HIV-1 adaptation at the population level. The differential adaptation of HIV-1 to MSM and HSX, which in most geographical regions show little inter-mixing, may have led over the years to the selection and, possibly, fixation of different adaptive mutations in the T/F viruses in the two groups. Genetic differences have been observed between T/F strains in MSM and HSX in small cohorts^3^.

Future studies may establish them at the population level, as sequencing technologies that allow facile identification of T/F viruses emerge. The technologies may also serve to elucidate such differences between other infected groups, which are likely to be present to lower degrees than between MSM and HSX, depending on the differences in the selection bias between the groups, the exclusivity of the associated modes of transmission, and the extent of mixing between the groups. Our findings also suggest that heritable viral traits such as SPVL^21^ may have evolved differently in MSM and HSX, potentially driving differential spread of the HIV-1 epidemic in the two groups. The extent of these differences may determine whether intervention strategies, including the development and use of preventive vaccines, may have to be tailored to individual infected groups.

## Methods

### Data of CD4 counts

To test our hypothesis that early CD4 counts in HSX would be higher than in MSM at the population level, we collated data from all large studies (*n* ≳ 1,000) that reported CD4 counts either at diagnosis or seroconversion in both these groups. The data are summarized along with our analysis in Table 1 and details are in Tables S3-S5. From reports on countries in the EU/EEA and China^10,13^, we digitized the median CD4 counts using WebPlotDigitizer (*https://automeris.io/WebPlotDigitizer*). For our analysis, we averaged the data over the study duration. To obtain sample sizes, we multiplied the diagnosed cases with the reported fraction of diagnoses contributing to the annual CD4 counts in the entire EU/EEA (Table S5). The fraction was assumed to be the same across the risk groups and the set of 21 countries studied. We also assumed the proportions of men and women in HSX to remain the same during 2010-18. In the CASCADE study^15^, which segregated data into age groups, we averaged over age groups. To obtain the population-weighted average CD4 counts, we assumed that the proportions of the populations in the different transmission categories were the same across age groups and that the fractions of men and women remained conserved (except in MSM and hemophiliacs) (Table S3). To calculate *R_T/F_*, we also collated data of CD4 counts from healthy, uninfected adults in the USA, UK, Italy (which was used for the three studies involving European populations), Tanzania, and China, which are listed in Table S6. For *R_T/F_* calculations pertaining to the UK, CD4 counts from healthy MSM and HSX were available, which we used. We found the counts in MSM comparable to those from healthy HSX men. As a result, for other populations, we used the cell counts for healthy HSX men where counts from healthy MSM were unavailable.

### Estimation of mean CD4 counts and their standard deviations

When the median, *m*, and interquartile range (IQR), (*q_l_, q_u_*), of CD4 counts were available, we estimated the corresponding mean, *μ*, and standard deviation (SD), *σ*, using 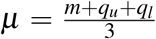 and 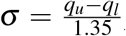, following the widely used method^22^ applicable to large sample sizes, as considered here. When 95% confidence intervals (CIs), (*c_l_, c_u_*), were available instead of IQR, we evaluated SD using another method^23^ which yielded 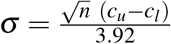 when the sample size *n* ≳ 100. When IQR was unavailable, we approximated the medians as the means, assuming the distributions to be normal. For data from China and EU/EEA, where *σ* was available for the total population, consisting of all the transmission categories, we estimated *σ* for MSM and HSX using the ratios between *σ* corresponding to MSM or HSX and the total population reported from other studies (see footnote in Table S4). Similarly, for obtaining *σ* for HSX men and women, we employed the corresponding ratios of maximum *σ* from the CASCADE study. When information necessary to estimate *σ* was unavailable, we used the highest *σ* available from the most relatable dataset, as with the UK and the CASCADE study. To estimate the SD of *R_T/F_*, we employed the error propagation equation^24^ and derived 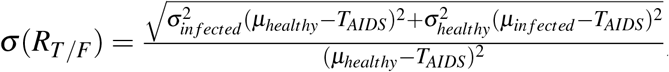, where *μ,σ* are given in Tables 1 and S6. For *σ*(*R_T/F_*) of all the data combined, we chose *σ* from EU/EEA, involving data from 21 countries.

### Estimation of CD4 counts at seroconversion

In the US study^14^, a model of CD4 count decline following seroconversion has been proposed, which allowed us to estimate CD4 counts at seroconversion from measurements at diagnosis. According to the model, the CD4 count *T* in an untreated individual at time *t* from seroconversion follows 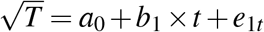, where *a*_0_ and *b*_1_ are constants and *e*_1*t*_ is an error term. At seroconversion, the CD4 count, *T*_0_, was obtained by setting *t* = 0, so that 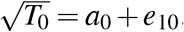. Assuming that *e*_1*t*_ = *e*_10_, it followed that 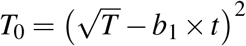. The values of *b*_1_ for different age groups and transmission categories were available^25^. Also, the median delays (and IQR) in diagnosis following seroconversion, *t_d_*, have been estimated^14^, using which we calculated the corresponding mean and SD. For MSM and HSX, we took the mid-value of the means of *t_d_* in 2006 and 2015 and chose the largest SD, and obtained *t_d_* = 4.05 *±* 6.67 and *t_d_* = 5.40 *±* 9.04 years, respectively, for the duration 2006–15. If *T_d_* is the CD4 count at diagnosis, then 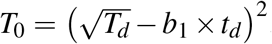. We applied the analysis to data from the most populated age group (13–29 years) and used the mid-value, 21 years, for which *b*_1_ was −0.93, −0.77, and −0.80 year^−1^for MSM, HSX men, and HSX women, respectively. Furthermore, we assumed the fractions of females to be the same among all non-MSM groups, in order to obtain the population sizes of MSM and HSX in this age group (footnote in Table S3). Correspondingly, we obtained *b*_1_ = −0.79 year^−1^for HSX. To obtain uncertainties in the estimates of *T*_0_, we repeated the above analysis with *T_d_* and *t_d_* set at values *±σ* away from their respective means, but ensuring that their lowerbounds *≥* 0 and omitting terms that are second order in *σ*. Half the difference between the resulting maximum and minimum values of *T*_0_ yielded the *σ* corresponding to seroconversion.

### Statistical analysis

To examine whether the mean CD4 counts (or mean *R_T/F_*) were significantly higher (or lower) in MSM than HSX, we employed the one-tailed t-test with unequal variance with the test statistic 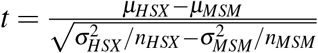 and degrees of freedom 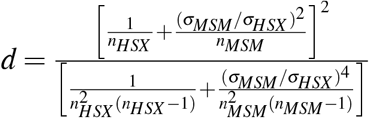, where *n_HSX_* and *n_MSM_* were the two sample sizes, respectively^26^. The tests were performed using the R package^27^, which yielded corresponding *P* values.

### Data of HIV-1 subtype prevalence

To assess the extent of mixing between MSM and HSX, we collated data of the prevalence of HIV-1 subtypes in the two groups across relevant geographical regions and calendar years. The data are summarized in Fig. 2 and Tables S1-S2 and discussed in Text S1.

### Data of association with transmission clusters

Finally, we considered the extent of association of MSM and HSX with transmission clusters as an indicator of the time of onward transmission post-infection. The corresponding data we collated along with data of the compositions of the largest transmission clusters in different settings are in Fig. 3 and Table S7 and are discussed in Text S2.

**Figure 3:**
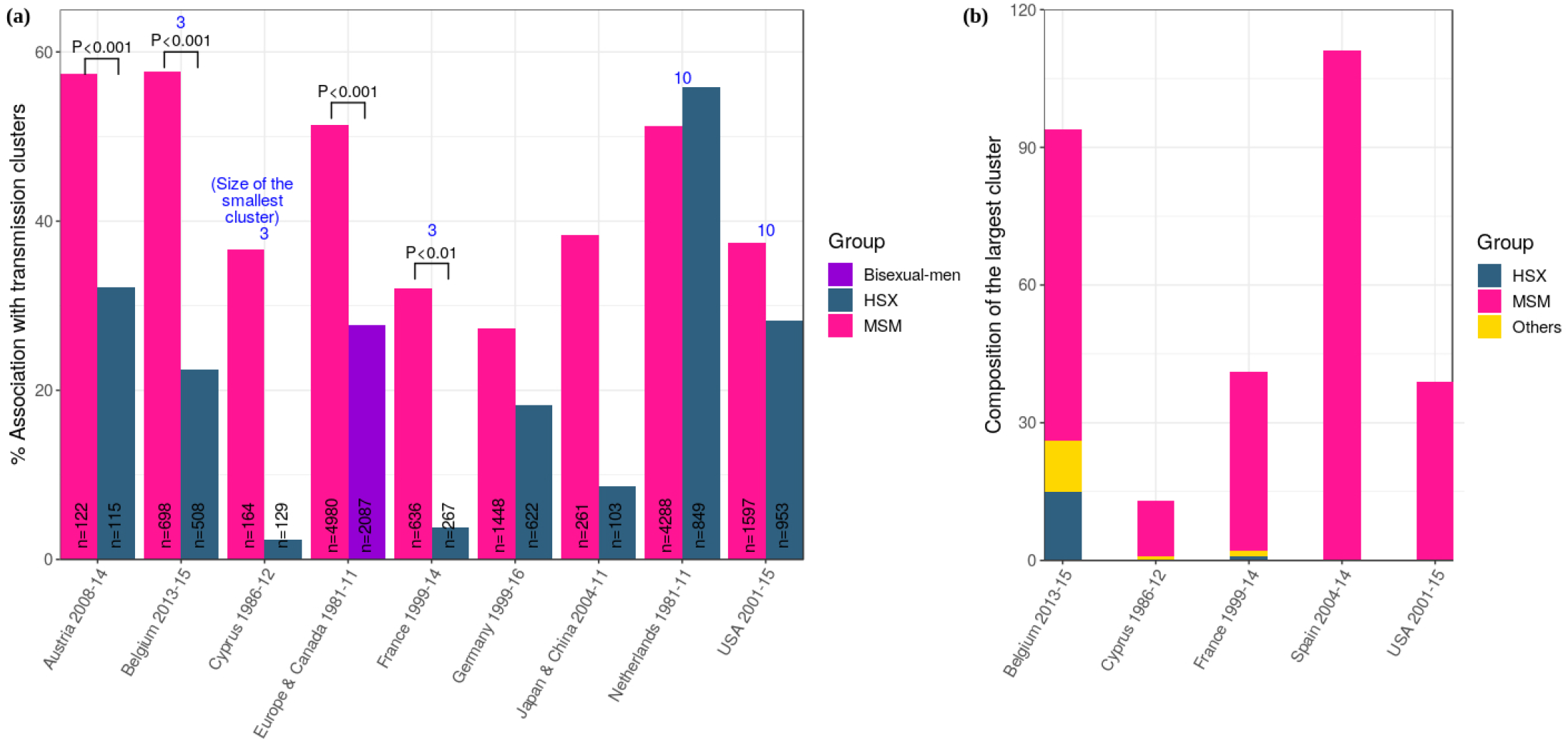
Association with transmission clusters. **(a)** The fraction of HSX and MSM (or bisexuals in one case) associated with transmission clusters and **(b)** the composition of the largest clusters in different geographical regions. The sample sizes (*n*) along with the time periods of the surveys are indicated. P values and the minimum sizes of the clusters (blue text) where available from the original sources are listed. Sources of the data and additional details are in Table S7. MSM are thus far more likely to be associated with clusters and also tend to form large clusters.

## Data Availability

All the data used in the study has been previously published. The sources are indicated in the manuscript. No new data is generated as part of this study.

## Acknowledgments

We thank Pranesh Padmanabhan, Rajat Desikan, and Pradeep Nagaraja for comments. This work was supported by the DBT/Wellcome Trust India Alliance Senior Fellowship IA/S/14/1/501307 (NMD).

## SUPPLEMENTARY INFORMATION

**Text S1: Distinct subtype prevalences indicate minimal mixing between MSM and HSX**

In many geographical locations, mixing between MSM and HSX appears minimal. This is evident from the different prevalences of HIV-1 subtypes in the two groups. MSM in western nations are dominated by HIV-1 subtype B, whereas HSX comprise a mixture of subtypes^1^, with subtypes B and C being the predominant ones^2^. For instance, in the United Kingdom, from 2002-2010, MSM had nearly 90% subtype B infections, whereas HSX had a little over 10% subtype B. Mixing between the two groups would have led to a more similar distribution of subtypes in the two. The two groups thus appear to have sustained their respective infections over the years in near complete isolation. The difference in subtype prevalences holds also in Canada, Spain, France, and other nations (Fig. 2(a); Table S1). In China, the dominant subtype is CRF01-AE, which is present in MSM with a frequency of >50% but in HSX at <40% (Fig. 2(b); Table S2)^3^, perhaps indicative of more mixing than in Europe. In Korea, the extent of mixing could not be assessed using subtypes because over 80% of all infections were subtype B^4^. In USA, though subtype B dominates both MSM and HSX^5,6^, mixing between the groups has been argued not to be common^7^. In the Nordic states, some mixing between MSM and HSX is evident^8^. Overall, little mixing between MSM and HSX is evident in most geographical settings, suggesting that the different selection biases between the groups may have been sustained over the course of the epidemic.

**Text S2: Clustering and transmission patterns**

MSM are known to engage in different sexual contact patterns compared to HSX. They tend to have more partners than HSX^9,10^. They are also far more likely to belong to transmission clusters compared to HSX^1^. A transmission cluster comprises individuals carrying viral genomes that cluster together in a phylogenetic tree^11^, suggesting that the viral sequences isolated from the individuals are closely related. In Japan and China, an infected MSM had a nearly 40% chance of being part of a cluster, whereas an infected HSX had <10% chance^12^. In France, the corresponding numbers were ∼35% and ∼4%, respectively^13^. This trend was true for all the countries with data available except the Netherlands (Fig. 3(a); Table S7). MSM also formed larger clusters than HSX. The largest clusters reported in Belgium and Spain comprised nearly 100 individuals each, with the Belgian cluster containing ∼70 MSM and the Spanish cluster exclusively MSM (Fig. 3(b); Table S7)^14,15^. Together, these data suggest greater similarity in the viral strains in MSM than HSX. One way in which this greater similarity could arise is by onward transmission occurring sooner after infection in MSM than HSX, allowing lesser individual host-specific adaptation before transmission.

### Supplementary Figures

**Figure S1.**
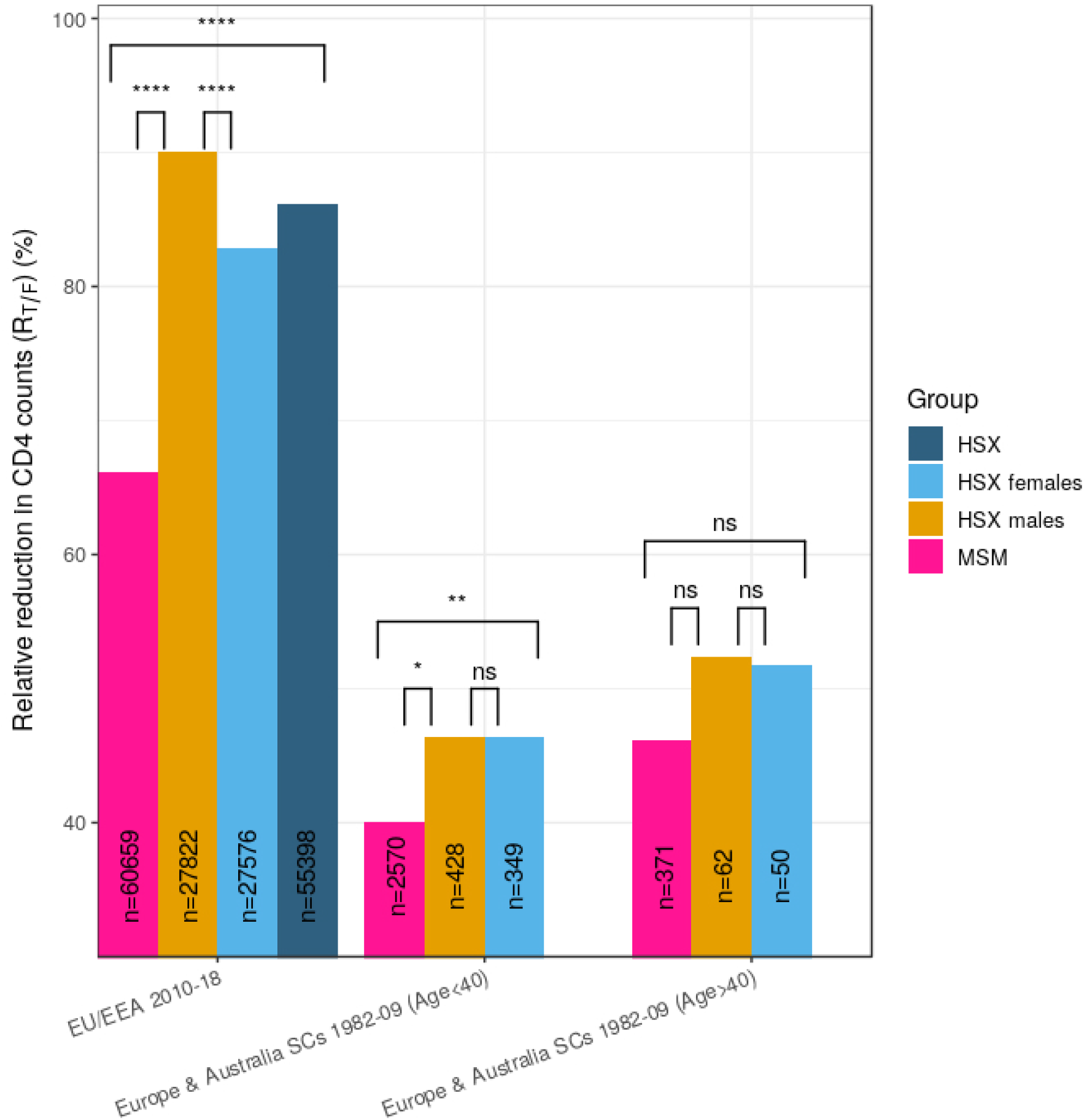
Effect of gender on the relative reduction in early CD4 counts. *R_T/F_* from EU/EEA during 2010–18^16^ and the CASCADE study^17^ indicate that HSX men have higher *R_T/F_* than MSM, with this difference achieving significance with large sample sizes, ruling out gender as a cause of the lower CD4 count reduction in MSM than HSX. The sample sizes (*n*) are indicated along with the *P* values. (****, ***, ** and * indicate P< 10^−4^, P< 10^−3^, P< 10^−2^and P< 0.05, respectively, while ns (not significant) implies P> 0.05.) Additional details are in Table 1 (main text).

### Supplementary Tables

**Table S1.** Prevalence of subtype B in Europe. Data from different regions in Europe show substantially higher subtype B percentage prevalence in MSM than HSX (*P* < 0.001 in each study, unless specified). The sample sizes (*n*) are in parantheses.

| Region<br>[Reference] | Survey<br>year(s) | Prevalence of subtype B (%) |  |
| --- | --- | --- | --- |
| | | MSM ($n$) | HSX ($n$) |
| France<sup>18</sup> | 2001 | 80.5 (113) | 53.2 (186) |
|  | 2006 – 07 | 90.6 (223) | 27.2 (202) |
|  | 2010 – 11 | 83.3 (311) | 26.2 (263) |
| Italy<sup>19</sup> | 2000 – 14 | 81.5 (1,824) | 54.9 (1,573) |
| UK<sup>20</sup> | 2002 – 10 | 88.4<sup>‡</sup> (21,321) | 13.3<sup>‡</sup> (38,948) |
| Madrid, Spain<sup>21</sup> | 2000 – 11 | 88.2 (340) | 41.5 (369) |
| Comunidad Valenciana (CV), Spain<sup>15</sup> | 2004 – 14 | 92.0<sup>‡</sup> (637) | 60.8<sup>‡</sup> (204) |
| Europe<sup>22</sup> | 2002 – 07 | 90.4 (2,084) | 33.5 (1,501) |
| Canada* & Europe<sup>23</sup> | 1998 – 10 | 95.5<sup>‡</sup> (5,570) | 38.7<sup>‡</sup> (2,052) |
| Japan<sup>12</sup> | 2004 – 11 | 96.9<sup>‡</sup> (261) | 39.8<sup>‡</sup> (103) |

<sup>‡</sup> $P$ value not specified.
\*In this study, 15.9% patients were from Canada.

**Table S2.** Prevalence of subtypes in China. A recent review^3^ of 130 published articles, together involving of 10,516 MSM and 6,759 HSX individuals, has examined the prevalence of different subtypes in China, which is reproduced below. *P* values indicate significant differences in the prevalences of 3 subtypes.

| Subtype | Prevalence (%) (95% CI) |  |  |
| --- | --- | --- | --- |
| | MSM | HSX | $P$ value |
| CRF01_AE | 51.28 (46.15 – 56.40) | 38.78 (33.08 – 44.63) | < 0.01 |
| CRF07_BC | 19.98 (16.17 – 24.07) | 14.88 (10.96 – 19.23) | 0.083 |
| CRF08_BC | 0.00 (0.00 – 0.00) | 9.81 (6.44 – 13.70) | < 0.01 |
| B\B' | 17.74 (12.78 – 23.26) | 15.41 (11.15 – 20.16) | 0.508 |
| C | 0.00 (0.00 – 0.00) | 1.89 (0.81 – 3.28) | < 0.01 |
| Others | 1.56 (0.98 – 2.24) | 2.39 (1.33 – 3.66) | 0.2109 |

**Table S3.**
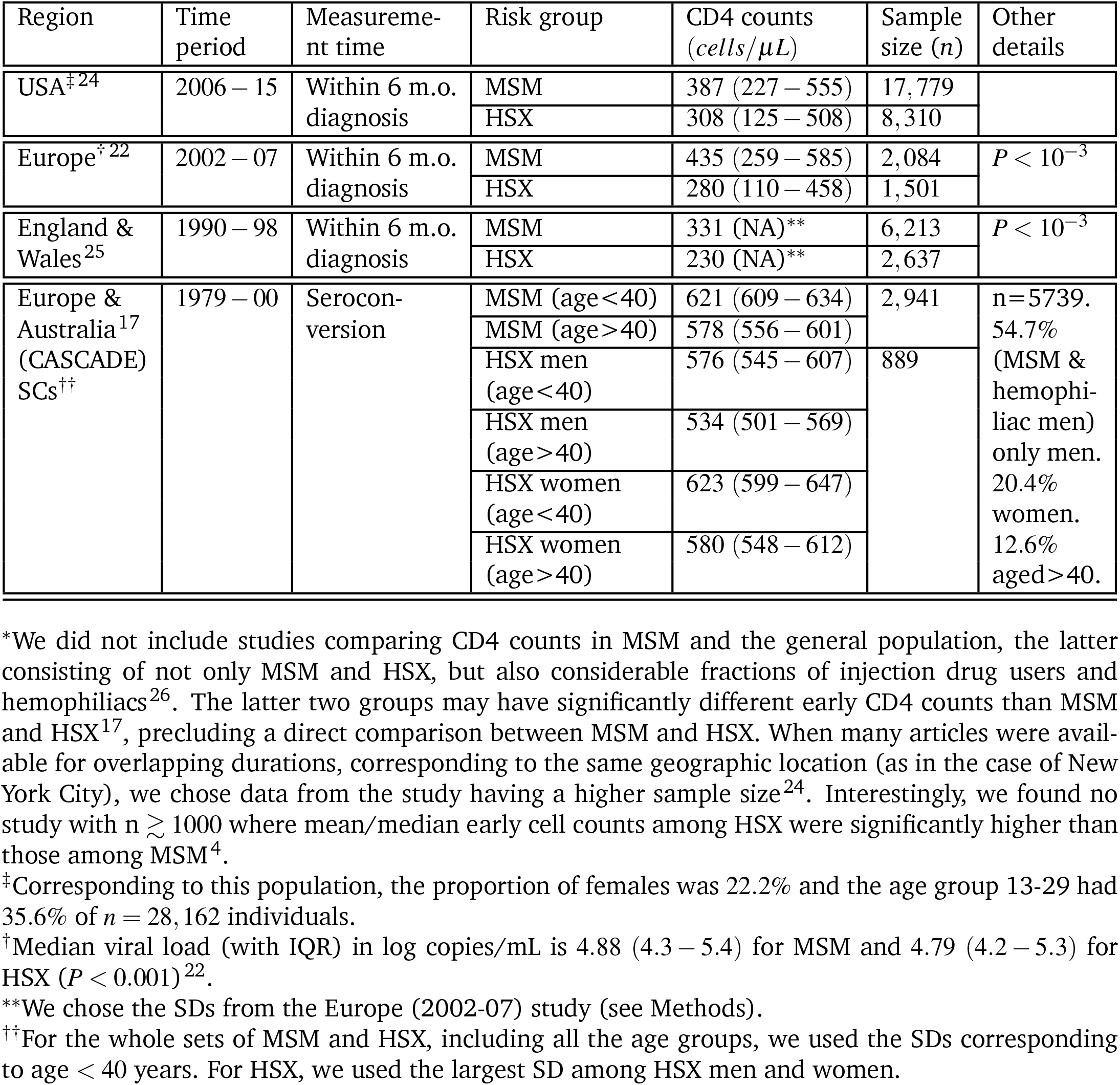
Early median CD4 cell counts in infected adults from several large population studies*. The sources of the studies, the periods of study, measurement times, and other details are mentioned. The USA and European studies report IQRs, whereas the CASCADE study provides 95% CIs. NA − not available.

**Table S4.**
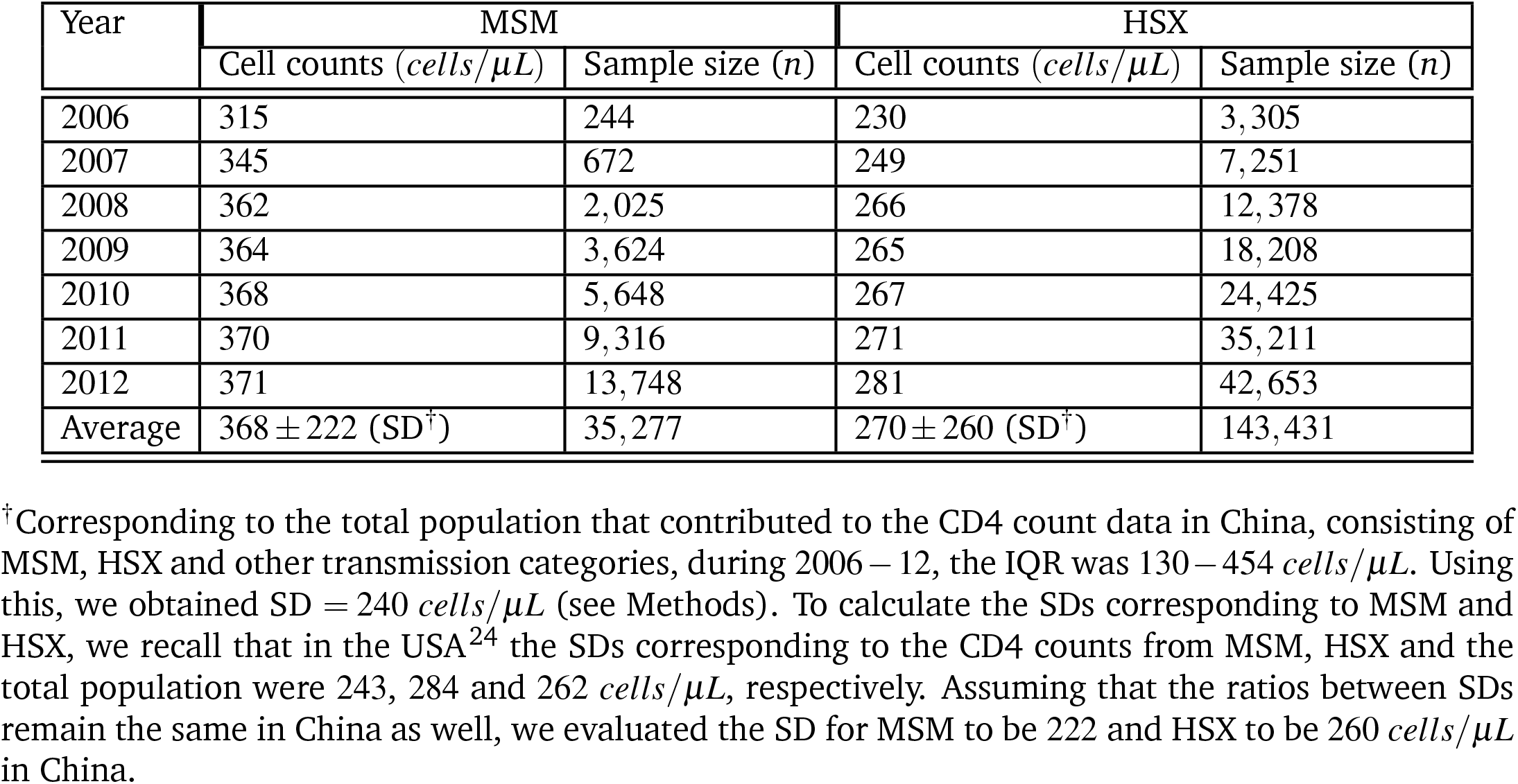
Early median CD4 cell counts in infected MSM and HSX from China^27^. The sample sizes (*n*) were directly available, while the cell counts were estimated using WebPlotDigitizer^28^. The last row represents estimates (see Methods) for the entire period 2006-12.

| Year | MSM |  | HSX |  |
| --- | --- | --- | --- | --- |
| | Cell counts (*cells/μL*) | Sample size ($n$) | Cell counts (*cells/μL*) | Sample size ($n$) |
| 2006 | 315 | 244 | 230 | 3,305 |
| 2007 | 345 | 672 | 249 | 7,251 |
| 2008 | 362 | 2,025 | 266 | 12,378 |
| 2009 | 364 | 3,624 | 265 | 18,208 |
| 2010 | 368 | 5,648 | 267 | 24,425 |
| 2011 | 370 | 9,316 | 271 | 35,211 |
| 2012 | 371 | 13,748 | 281 | 42,653 |
| Average | $368 \pm 222$ (SD<sup>†</sup>) | 35,277 | $270 \pm 260$ (SD<sup>†</sup>) | 143,431 |

<sup>†</sup>Corresponding to the total population that contributed to the CD4 count data in China, consisting of MSM, HSX and other transmission categories, during 2006 – 12, the IQR was 130 – 454 *cells/μL*. Using this, we obtained SD = 240 *cells/μL* (see Methods). To calculate the SDs corresponding to MSM and HSX, we recall that in the USA<sup>24</sup> the SDs corresponding to the CD4 counts from MSM, HSX and the total population were 243, 284 and 262 *cells/μL*, respectively. Assuming that the ratios between SDs remain the same in China as well, we evaluated the SD for MSM to be 222 and HSX to be 260 *cells/μL* in China.

**Table S5.** Early median CD4 cell counts in infected adults from EU/EEA^16^. The cell counts were estimated using WebPlotDigitizer^28,29^. The median ages, whenever available, are also provided. The last row provides the mean cell counts (with SDs) and total numbers of MSM, HSX men and women, respectively, estimated as in Methods.

| Year | No. of diagnoses* |  | Fraction with CD4 counts<sup>††</sup> | Median age |  | Median CD4 count (*cells/μL*) |  |  |
| --- | --- | --- | --- | --- | --- | --- | --- | --- |
|  | MSM | HSX |  | MSM | HSX | MSM | HSX men | HSX women |
| 2010 | 10,348 | 11,693 | 0.591 | - | - | 420 | 261 | 312 |
| 2011 | 10,411 | 11,011 | 0.561 | - | - | 422 | 260 | 317 |
| 2012 | 11,238 | 10,782 | 0.553 | - | - | 436 | 270 | 319 |
| 2013 | 11,553 | 10,187 | 0.607 | - | - | 443 | 271 | 339 |
| 2014 | 11,821 | 10,049 | 0.614 | - | - | 450 | 281 | 346 |
| 2015 | 11,484 | 9,267 | 0.747 | - | - | 454 | 284 | 345 |
| 2016 | 10,508 | 9,003 | 0.673 | 34 | 39 | 440 | 274 | 351 |
| 2017 | 9,723 | 8,518 | 0.717 | 34 | 39 | 434 | 268 | 349 |
| 2018 | 8,049 | 7,267 | 0.695 | 36 | 41 | 423 | 262 | 340 |
| Total | $n_{MSM}^{**} = 60,659, n_{HSXM}^{**} = 27,822, n_{HSXW}^{**} = 27,576$ | | | | | $(437 \pm 397)^{\ddagger}$ | $(270 \pm 464)^{\ddagger}$ | $(335 \pm 325)^{\ddagger}$ |

\*The number of diagnoses specific to each country during 2009-18 was provided for MSM and HSX in the latest annual report<sup>16</sup>. Using these, we estimated the numbers above.
††These are taken from the annual surveillance reports<sup>29</sup>. Since this number was unavailable for 2009, we omitted the data for the calculation of the mean CD4 counts for the combined data.
\*\*The numbers of HSX men and women diagnosed from the 21 EU/EEA countries in 2018 were available in the 2019 annual report<sup>16</sup>. Using this, we calculated the fractions of HSX men and women and assumed that they remained the same during 2010-18. Also, $n_{HSX} = n_{HSXmen} + n_{HSXwomen} = 55,398$ . (There were a small number of transgenders among HSX in column 3, but the CD4 counts were reported only for HSX men and women.)
‡In 2018, for 16,694 adults across risk groups, the median cell counts with 95% CIs were 365 (359 – 372) *cells/μL*. Using this, we computed their SD to be $\sim 429$ *cells/μL* and evaluated the SDs for MSM and HSX. To obtain the SDs for HSX men and women, we used the SD ratio for the age group < 40 years corresponding to the CASCADE study (see Methods).

**Table S6.** CD4 T cell counts in healthy adults. Mean CD4 counts in healthy adults from different population groups which define baseline counts for estimating the relative reduction in early cell count following HIV-1 infection. Sample sizes are in brackets. SD is standard deviation.

| Region | Category<br>(sample size, *n*) | Cell counts<br>(*cells/μL*) | Other details |
| --- | --- | --- | --- |
| China<sup>30,31</sup><br>(Hong Kong<sup>30</sup> & Shanghai<sup>31</sup>) | Men (78<sup>30</sup> & 377<sup>31</sup>) | 725 | Combined mean; SD = 258<sup>30</sup> & 256<sup>31</sup>. |
|  | Women (130<sup>30</sup> & 237<sup>31</sup>) | 724 | Combined mean; SD = 254<sup>30</sup> & 255<sup>31</sup>. |
|  | HSX (822) | 725 | SD = 255 for Shanghai<sup>31</sup> |
| Sub-Saharan Africa<br>(Tanzania)<sup>32</sup> | Men* (42) | 666 | SD = 247 |
|  | Women* (60) | 802 | SD = 250 |
|  | HSX (102) | 746 | SD = 257<sup>†‡</sup> |
| USA<sup>33</sup> | Men** (33) | 921 | SD = 188 |
|  | Women (67) | 1041 | SD = 340 |
|  | HSX** (100) | 1001 | SD = 305 |
| UK<sup>34</sup> | HSX men<sup>†</sup> (50) | 840 | SD = 285 |
|  | HSX women (50) | 1050 | SD = 377 |
|  | HSX<sup>‡</sup> (100) | 945 | We used the SD for women. |
|  | MSM<sup>†,‡</sup> (100) | 800 | SD = 324 |
| EU/EEA (& Europe) & Australia<br>(Italy)<sup>35</sup> | General population or HSX<sup>††</sup> (965) | 941 | SDs were not reported. We used the SDs from the UK study (see Methods). |
|  | Men<sup>††</sup> (532) | 902 |  |
|  | Women (436) | 989 |  |

\* $P = 8 \times 10^{-3}$ for the comparison between men and women (reported in the original study<sup>32</sup>).
\*\* $P = 0.04$ for the comparison between healthy men and HSX from USA.
<sup>†</sup> $P = 0.22$ for the comparison between MSM and HSX men from UK.
<sup>‡</sup> $P = 6.6 \times 10^{-4}$ for the comparison between healthy MSM and HSX from UK, using SD (= 305) for HSX from USA. Using SD (=377) corresponding to HSX women from UK instead yielded $P = 2 \times 10^{-3}$ .
<sup>††</sup> $P = 0.018$ using the SDs from UK.
<sup>‡‡</sup>For our calculation involving the EU/EEA 2010-18 population (footnote in Table 1), we used the SD (=377) for the HSX from Europe/UK.

**Table S7.** Association of MSM and HSX with transmission clusters. The percentages of individuals found to be associated with transmission clusters in MSM and HSX in several studies are collated. In the second column, the numbers in parantheses indicate sample sizes examined. Where available, P values and the largest cluster sizes are indicated in other details.

| Region<br>(Study period) | Group (n) | Associated with<br>clusters (%) | Other details |
| --- | --- | --- | --- |
| Japan<sup>12</sup><br>(2004 – 11) | MSM (261) | 38.3 | Clusters are also linked to China. |
|  | HSX (103) | 8.7 |  |
| Austria<sup>36</sup><br>(2008 – 14) | MSM (122) | 57.4 | $P < 0.001$ |
|  | HSX (115) | 32.2 |  |
| Belgium<sup>14</sup><br>(2013 – 15) | MSM (698) | 57.7 | $P < 0.001$. Cluster size $\geq 3$.<br>The largest cluster is sized 94,<br>consisting of 68 MSM and 15 HSX. |
|  | HSX (508) | 22.4 |  |
| Cyprus<sup>37</sup><br>(1986 – 2012) | MSM (164) | 36.6 | Cluster size $\geq 3$. The two largest clusters<br>are sized 13, having 11 – 12 MSM. |
|  | HSX (129) | 2.3 |  |
| France<sup>13</sup><br>(1999 – 2014) | MSM (636) | 32.1 | $P < 0.01$. Cluster size $\geq 3$.<br>The largest cluster, sized 41,<br>has 39 MSM and a HSX. |
|  | HSX (267) | 3.8 |  |
| Germany<sup>38</sup><br>(1999 – 2016) | MSM (1,448) | 27.3 |  |
|  | HSX (622) | 18.2 |  |
| Netherlands<sup>39</sup><br>(1981 – 2011) | MSM (4,288) | 51.2 | Cluster size $\geq 10$. |
|  | HSX (849) | 55.8 |  |
| Spain<sup>15</sup><br>(2004 – 14) | MSM (637) | – | 8 of 12 clusters sized $\geq 10$ consist only<br>of MSM. The largest cluster has a size<br>of 111, and is composed only of MSM. |
|  | HSX (204) | – |  |
| USA<sup>40</sup><br>(2001 – 15) | MSM (1,597) | 37.4 | The largest cluster has a size<br>of 39, and has only MSM. |
|  | HSX (953) | 28.3 |  |
| Canada and<br>9 European<br>countries<sup>41</sup><br>(1981 – 2011) | MSM (4,980) | 51.4 | The 9 countries are Italy, Greece,<br>Netherlands, Germany, France,<br>UK, Norway, Austria, and Spain.<br>$P < 0.001$. |
|  | Bisexual<br>men (2,087) | 27.7 |  |

